# The Effect of Bilingual Exposure on Language and Cognitive Development in Children Following Ischemic Stroke

**DOI:** 10.1101/2021.10.27.21265481

**Authors:** Kai Ian Leung, Nomazulu Dlamini, Robyn Westmacott, Monika Molnar

## Abstract

**AIM:** While many children who experience ischemic stroke come from bilingual backgrounds, it is unclear whether bilingual exposure affects post-stroke development. Our research evaluates the effects of bilingual vs. monolingual exposure on linguistic/cognitive development post-stroke.

**METHOD:** An institutional stroke registry and medical charts were used to gather data on 237 children across 3 stroke-onset groups: neonatal - <28 days, first-year - 28 days to 12 months and childhood - 13 months to 18 years. The Pediatric Stroke Outcome Measure (PSOM) was administered at several times post-stroke, to evaluate cognition and linguistic development.

**RESULTS:** Bilingual children had better post-stroke performance on the language subscales, compared to monolinguals. An interaction with stroke-onset group was also observed, with monolinguals in the first-year group having worse outcomes.

**INTERPRETATION:** Overall, no detrimental effects of bilingualism were found on children’s post-stroke cognition and linguistic development. Our study suggests that a bilingual environment may facilitate language development in children post-stroke.

## Introduction

Pediatric stroke is caused by a blockage or a rupture of the blood vessels of the brain and differs from adult stroke in clinical presentation, etiology, risk factors and outcomes^1^. More than half of all strokes in children are arterial ischemic stroke (AIS), making up 70% of all ischemic strokes in children^1^. AIS is due to an interruption of blood flow to the brain, in an arterial distribution secondary to occlusion of cerebral arteries^2^. AIS most often presents as a focal neurologic deficit occurring in 61% of cases^3^. There are important differences in etiology and outcome based on child’s age at stroke onset, which is typically categorized into neonatal stroke (28 weeks’ gestation to 28 postnatal days of life) or childhood stroke (infancy to 18 years of age)^4^.

Cognitive and language development is often negatively affected by AIS. When most domains of language are considered (including phonology, semantics, syntax, discourse, literacy), children with strokes during the neonatal period up to 1^st^ year of life have worse language outcomes compared to childhood stroke^5^. Lesion laterality, or the affected hemisphere of the stroke, does not seem to affect language development after neonatal stroke^6^, while in childhood stroke, it has been found to lead to language impairments after left hemisphere damage similarly to adults^7^. Younger age at stroke (before 1st year) was also found to be a predictor of worse cognitive outcomes^8,9^, particularly when response inhibition, divided attention, switching, sustained attention and other executive function evaluations are considered^10–12^. Similar to language, the effect of lesion laterality on cognitive outcomes has proven inconsistent^11^; risk factors for worse cognitive outcome include larger lesion volume^10^, infarcts affecting cortical and subcortical regions^8^ and those involving both small and large vessel territories^13^.

Research concerning the effects of specific demographic and psychosocial factors on cognitive and behavioral outcomes in pediatric stroke are emerging^14^, however an unexplored area remains the socio-linguistic factor of bilingualism. Bilinguals, or those who are learning and using two or more languages, make up half of the world’s population^15^. While a prior belief was that learning two languages would have a negative impact on development, recent research demonstrates that neurotypical monolingual and bilingual children reach the developmental milestones of language and cognitive development at the same pace^16^. Moreover, it has been proposed that the bilingual environment might afford certain benefits cognitively and linguistically^17^. According to the Bilingual Cognitive Advantage hypothesis, the mental exercise related to using two languages (e.g., activating the appropriate language, switching between languages, etc.) can positively affect non-linguistic *cognitive* functions^18^. Recent meta-analyses support the existence of a bilingual advantage in executive functions in neurotypical pediatric populations^19^. Childhood bilingualism is also associated with *linguistic* advantages. Specifically, considering metalinguistic abilities, bilinguals tend to consistently outperform their monolingual peers^20^. Bilingualism may also facilitate language production skills, such as novel word learning in bilingual school-aged children^21^.While there is an ongoing debate about the existence of bilingual advantage, the current work intends to move beyond the dichotomy of advantage vs. no advantage and focus on how bilingual exposure may interact with (cognitive and/or language) development in populations with neurologic deficits, such as ones caused by AIS.

### The Current Study

Here, we investigate whether bilingual vs. monolingual exposure affects *language* or *cognitive* development differently in children following ischemic stroke. This is important, as speaking two languages at home and having low English-proficiency parents are associated with adverse child health outcomes^22^; limited English-proficiency pediatric patients also experienced care-related disparities associated to their language use^23^. Research concerning bilingual exposure in pediatric stroke population and guidelines available to clinicians are scarce. Thus far, the only study that examined bilingual pediatric stroke populations did not include a control group of monolinguals^24^. Therefore, to date, we are not aware of any studies evaluating bilingual exposure on stroke recovery in children.

The specific objective of this study is to determine if bilingual exposure affects cognitive and linguistic development in children following stroke as measured by the Pediatric Stroke Outcome Measure (PSOM). Currently, the PSOM is the only available standardized neurological examination of outcome, specific to pediatric stroke recovery^25^. It is a reliable and valid composite outcome measure based on motor, language, and cognitive/behavioural dimensions, evaluated against standardized neuropsychological measures in each domain, and has been utilized in various research^12,13,26^. The PSOM scores analyzed in the current study were collected at hospital discharge and in the follow-up clinic, on average at 3-6 months and 12 months post-stroke, then at 1-2-year intervals after the first year up to 18 years of age. In our current sample, follow-up was at minimum 2 months post-stroke, up to a maximum 10.5-years post-stroke.

Given that bilingual exposure during childhood is not associated with any negative outcomes in terms of linguistic and cognitive development in neurotypical children, we hypothesized that bilingual exposure in pediatric stroke patients would not have any negative consequences. Accordingly, we predicted that the PSOM follow-up scores of bilingual children would not be worse than the scores of their monolingual peers. Further, considering the cognitive and linguistic advantages observed in bilinguals, it is a possibility that bilingual patients would benefit from their bilingual experience and show an advantage in their development, reflected through differences in performance in certain PSOM subscales (e.g., language and cognitive/behavioural). Finally, we predicted no differences between monolinguals and bilinguals if they suffered stroke within the neonatal period. This is because post-stroke outcome measures show consistent decline across populations post neonatal stroke^9^.

## Methods

### Patients

Patients were enrolled in the Hospital for Sick Children (SickKids; Toronto, Canada) Stroke Registry between January 1, 2008 and December 1, 2019, and were considered for the study if they were first-ever arterial ischemic stroke (AIS) patients aged 0-17 years at stroke. Exclusion criteria included: 1) presumed perinatal AIS; 2) multiple diagnoses of stroke (e.g., stroke recurrence or cerebral sinovenous thrombosis followed by AIS); 3) congenital/developmental disease or premorbid learning/psychological diagnosis prior to stroke onset, associated with language or cognitive outcomes (i.e., trisomy 21; nonverbal autism); 4) fewer than two PSOM evaluation scores available post-stroke. This study was approved by the Research Ethics Board at SickKids (REB#1000067719) and consent was provided for all participants or their caregivers as part of their enrollment in the SickKids Stroke Registry.

Outcome measure data was retrieved from the SickKids Stroke Registry. First and second language (and other languages, if applicable) and any described use or exposure were abstracted from medical charts. The monolingual group comprised of children who were exposed to one language only from birth. The bilingual group was defined by concurrent learning or exposure to at least two languages (e.g., one heritage/home language and one societal language). Children who began to learn a second language in school settings, a typical experience for sequential bilinguals, were subsumed into the bilingual group. Given that both simultaneous and sequential bilinguals have been associated with cognitive and linguistic advantages, both types of bilinguals were grouped together for analysis^27,28^.

Previous literature evaluating outcomes using the PSOM indicated that emerging deficits were more common in recovery patterns of children younger than 1 year at stroke onset^13^. Thus, patients were stratified into three ‘age at stroke’ groups as follows: neonates – <28 days; first-year – 28 days to 12 months; and childhood – 13 months to 18 years. Using Kruskal-Wallis test, our preliminary analyses evaluating this grouping also corroborated these differences using the Pediatric Stroke Outcome Measure, *H(2)=12*.*15, p<*.*002*, indicating a significant difference between the three groups.

### Pediatric Stroke Outcome Measure (PSOM)

The PSOM is a standardized, structured pediatric neurological examination comprised of 115 test items and is summarised into five subscales; Right Sensorimotor, Left Sensorimotor, Language Expression and Language Comprehension and Cognitive/Behavioural subscales^25,29^. Each PSOM subscale is scored on a range from 0 (no deficit), 0.5 (mild deficit), 1 (moderate deficit) to 2 (severe deficit), with a total PSOM out of 10 considering the sum of all 5 subscales. A greater score is indicative of worse outcome. The PSOM takes into consideration the administering neurologist’s observations (partially based on age appropriate language and cognitive tasks), parent’s report and medical chart information (e.g., clinic notes, neuropsychological assessment – including standardized language and cognition measures). The PSOM’s construct validity and reliability have been established against standardized neuropsychological measures, evaluating the content or domains matched to each subscale^25^.

### Statistical analysis

Anonymized data is available openly at https://doi.org/10.5683/SP2/E3ECVD and was analyzed in Jamovi (version 1.6.6, The jamovi project, 2020, https://www.jamovi.org/). Growth curve modelling, a mixed effects model including timepoint (month at test from stroke onset) as a predictor, was used to analyse the relationship between language group and outcome^30,31^. In our analyses, we focused on the Cognitive/Behavioural, Language Expression (i.e., language production) and Language Comprehension (i.e., language reception) PSOM subscales. A Combined Language subscale was calculated through sum of the scores from the language expression and comprehension subscales. We fit linear mixed models for each PSOM subscale as outcome variables, using timepoint (month at test from stroke onset), language group (monolingual, bilingual), age at stroke onset (neonate, first-year, childhood), sex (female, male), laterality (right, left, bilateral) and its interaction as fixed effects (see Supplementary Materials for full models and additional results related to sex and laterality, which are generally in line with findings reported in previous literature).

Here, we report models relevant to our hypothesis. These models include *timepoint* (month at test from stroke onset; each child was evaluated at multiple time points after stroke onset, up to 10 years), *language group* (monolingual vs. bilingual status), *age at stroke* (neonate, first-year, childhood) and its interactions. Timepoint variable was assigned a specified loading in the model, based on the month after stroke at test and was also used as random variable when fit improved. Maximum Likelihood method of estimation was used, and model fit was evaluated using Akaike Information Criterion and Bayesian Information Criterion values. Statistical analyses and reporting have been done in consultation with an institutional statistician.

## Results

### Study Cohort

253 children with AIS met our inclusion and exclusion criteria. Of these patients, three additional patients were excluded due to chart unavailability; 13 deceased patients had residual data in the registry and were excluded. The final study sample totalled 237 children. The demographic and stroke characteristics of all children are available in Table 1. The sample consisted of monolingual (n=111) and bilingual patients (n=126). This distribution is in line with Statistics Canada’s (2016) reports of 4 in 10 households speaking more than one language at home, with this proportion increasing for larger metropolitan areas^32^. While the language group distribution is fairly comparable in the neonate and first-year age at stroke groups, the childhood group had more monolinguals than bilinguals. Also, the first-year group had the least number of patients (only 14%) compared to the other groups. Socioeconomic status (SES) was available for a subset of the sample and was evaluated by calculating a combined SES score based on annual income and parents’ highest level of education. A Mann-Whitney test found no difference between monolinguals and bilinguals in socioeconomic status (U=867.50, p=0.65).

**Table 1.**
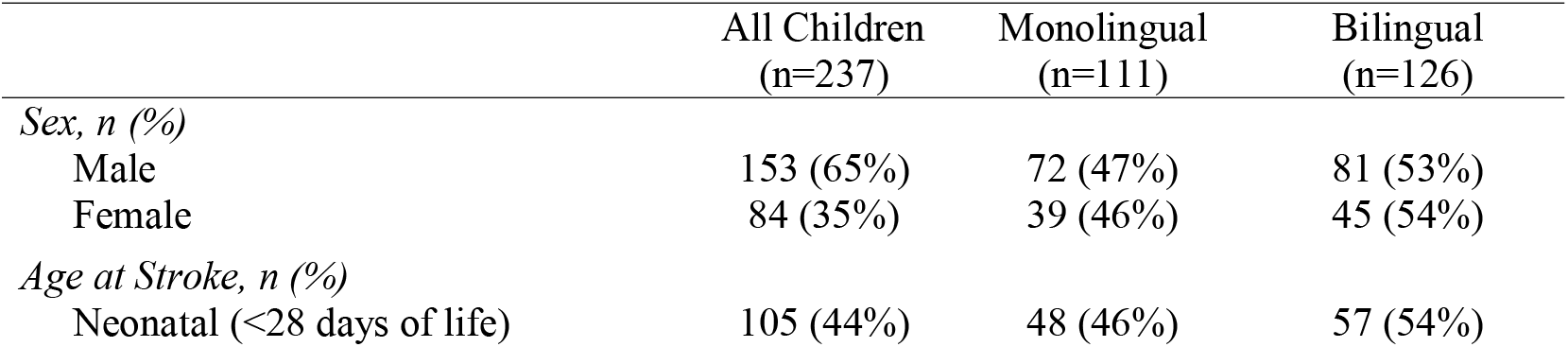
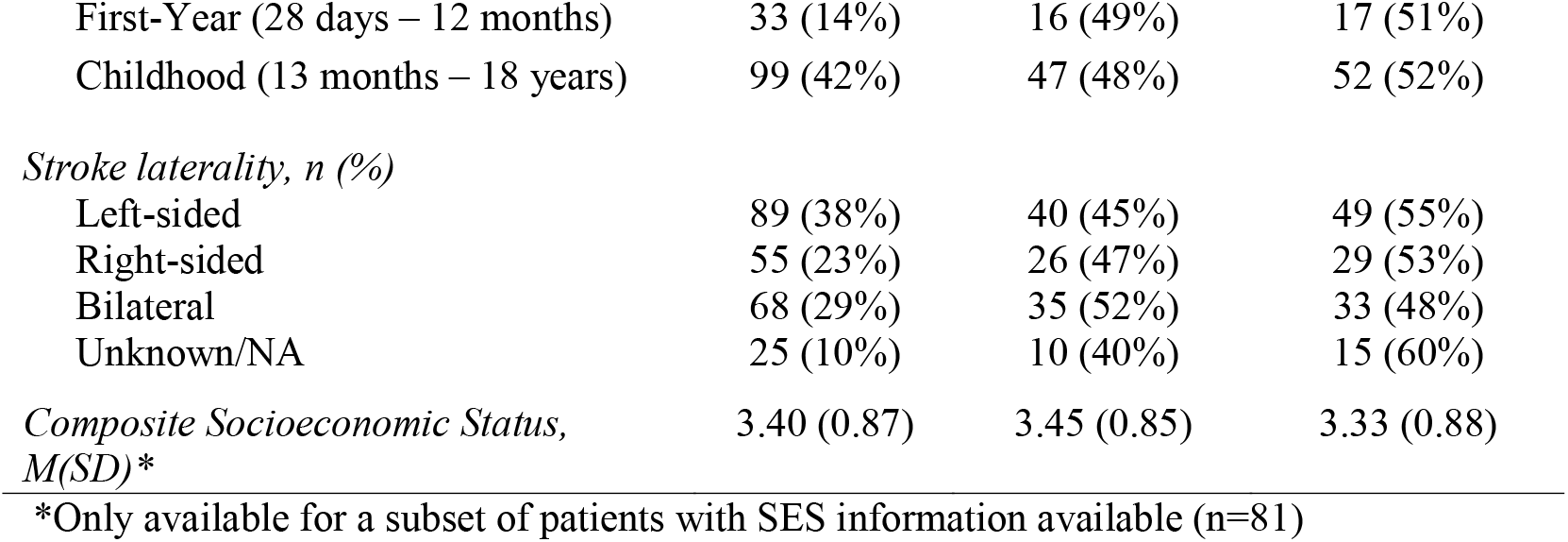
Patient demographics (sex, age at stroke group subdivided into 3 groups based on literature and preliminary analyses, socioeconomic status) and stroke characteristics (lesion laterality) for monolingual and bilingual participants

### Cognitive/Behavioural PSOM

We modelled cognitive development as predicted by language group, age at stroke group and its interaction, with timepoint variable added in as an additional fixed variable and random slope to increase model fit. While language group was nonsignificant, age at stroke was a significant predictor of cognitive/behavioural PSOM scores, such that PSOM was seen to increase incrementally with an older age at stroke indicating a worse outcome: the neonatal group had the lowest PSOMs, while the childhood group had highest PSOMs (Table 2).

**Table 2.** Summary of results from two one-sided tests (TOST) with independent variable of language group, on cognitive/behavioural PSOM subscale.

|  |  | Equivalence<br>Bounds:<br>90% CI | t | df | p |
| --- | --- | --- | --- | --- | --- |
| Cognitive | t-test |  | 1.91 | 718 | 0.056 |
| Behavioural | TOST Upper | 0.127 | -5.09 | 718 | <.001 |
| PSOM | TOST Lower | 0.00957 | 8.92 | 718 | <.001 |

#### Equivalence Testing of Language Group

As nonsignificant result is insufficient to conclude a lack of effect, further equivalence test analyses evaluating the effect of language group on the cognitive/behavioural PSOM performance were conducted. Two one-sided tests (TOST) independent samples t-test procedure was employed to investigate whether the effect size found is smaller than the smallest effect size of interest and aims to reject a null effect. Summary statistics are available in Table 2. For cognitive/behavioural PSOM, the equivalence test was significant, t(718)= 1.91, p = 0.056, given equivalence bounds of 0.009 and 0.127, suggesting no difference between groups on cognitive/behavioural PSOM.

### Combined Language Subscales PSOM

We modelled language development as predicted by language group, age at stroke group and its interaction, with timepoint variable as additional fixed effect as it did not increase fit as random effect. Language group was significant, with better outcomes favouring bilinguals over monolinguals (p=.04; Table 3). Additionally, one of the interaction comparisons between the age at stroke group and language group, showed significance (p=.04; Table 3), with better outcomes for bilinguals in the first-year group. Differing from the age at stroke group pattern apparent in the cognitive PSOM model, a U-shaped pattern with the highest scores observed for the first-year group.

**Table 3.** Summary of results from growth curve models predicting the fixed effects of timepoint, language group, age at stroke group and its interaction on various PSOM subscales, significant values are bolded.

| **Cognitive/Behavioural PSOM - Predictor** | **Estimate** | **SE** | **95% CI** | **p** |
| --- | --- | --- | --- | --- |
| (Intercept) | 0.2878 | 0.0261 | 0.2367-0.3390 | <.001 |
| Timepoint | 0.0033 | 7.08e-4 | 0.0020-0.0047 | <.001*** |
| Language group |  |  |  |  |
| Bilingual – Monolingual | -0.0705 | 0.0520 | -0.1723-0.0313 | 0.176 |
| Age at stroke group |  |  |  |  |
| First-year – Neonate | 0.1859 | 0.0694 | 0.0499-0.3219 | 0.008** |
| Childhood - Neonate | 0.3332 | 0.0498 | 0.2245-0.4198 | <.001*** |
| Language group × Age at stroke group | | | | |
| Bilingual – Monolingual × | -0.1088 | 0.1388 | -0.3808-0.1631 | 0.434 |
| First-year – Neonate |  |  |  |  |
| Bilingual – Monolingual × | -0.0190 | 0.0997 | -0.2144-0.1763 | 0.849 |
| Childhood - Neonate |  |  |  |  |
| **Combined Language PSOM - Predictor** | **Estimate** | **SE** | **95% CI** | **p** |
| (Intercept) | 0.362 | 0.0435 | 0.2765-0.4469 | <.001 |
| Timepoint | 3.43e-4 | 8.63e-4 | -0.0013-0.0020 | 0.691 |
| Language group |  |  |  |  |
| Bilingual – Monolingual | -0.178 | 0.0869 | -0.3479- -0.0073 | 0.042* |
| Age at stroke group |  |  |  |  |
| First-year – Neonate | 0.319 | 0.1161 | 0.0912-0.5461 | 0.007** |
| Childhood - Neonate | 0.111 | 0.0829 | -0.0519-0.2730 | 0.184 |
| Language group × Age at stroke group | | | | |
| Bilingual – Monolingual × | -0.464 | 0.2321 | -0.9186- -0.0088 | 0.047* |
| First-year – Neonate |  |  |  |  |
| Bilingual – Monolingual × | -0.108 | 0.1658 | -0.4326-0.2174 | 0.517 |
| Childhood - Neonate |  |  |  |  |
| **Language Expression PSOM - Predictor** | **Estimate** | **SE** | **95% CI** | **p** |
| (Intercept) | 0.2314 | 0.0260 | 0.1805-0.2822 | <.001 |
| Timepoint | 1.83e-4 | 5.21e-4 | -8.38e-4-0.0012 | 0.725 |
| Language group |  |  |  |  |
| Bilingual – Monolingual | -0.1170 | 0.0519 | -0.2187- -0.0152 | 0.025* |
| Age at stroke group |  |  |  |  |
| First-year – Neonate | 0.1804 | 0.0694 | 0.0445-0.3163 | 0.010** |
| Childhood - Neonate | 0.0431 | 0.0494 | -0.0537-0.1399 | 0.383 |
| Language group × Age at stroke group | | | | |
| Bilingual – Monolingual × | -0.3289 | 0.1387 | -0.6008- -0.0570 | 0.019* |
| First-year – Neonate |  |  |  |  |
| Bilingual – Monolingual × | -0.0605 | 0.0988 | -0.2541-0.1331 | 0.541 |
| Childhood - Neonate |  |  |  |  |
| **Language Comprehension PSOM - Predictor** | **Estimate** | **SE** | **95% CI** | **p** |
| (Intercept) | 0.1364 | 0.0201 | 0.0970-0.1757 | <.001 |
| Timepoint | 1.75e-4 | 4.30e-4 | -6.67e-4-0.0010 | 0.684 |
| Language group |  |  |  |  |
| Bilingual – Monolingual | -0.0671 | 0.0402 | -0.1459-0.0116 | 0.097† |
| Age at stroke group |  |  |  |  |
| First-year – Neonate | 0.1490 | 0.0537 | 0.0437-0.2543 | 0.006** |
| Childhood - Neonate | 0.0646 | 0.0384 | -0.0106-0.1398 | 0.094† |
| Language group × Age at stroke group |  |  |  |  |
| Bilingual – Monolingual × | -0.1578 | 0.1075 | -0.3685-0.0528 | 0.144 |
| First-year – Neonate |  |  |  |  |
| Bilingual – Monolingual × | -0.0534 | 0.0768 | -0.2038-0.0970 | 0.488 |
| Childhood - Neonate |  |  |  |  |

Note: † p < .10; \* p < .05; \*\* p < .01; \*\*\* p < .001

#### Language Comprehension PSOM

We modelled language comprehension as predicted by language group, age at stroke group and its interaction, with the timepoint variable as an fixed effect as model fit did not increase as a random effect. A trend for language group favoured better outcomes for bilinguals over monolinguals (p=.08; Table 3). As for age at stroke group findings, an U-shaped pattern with the highest scores was observed for the first-year group.

#### Language Expression PSOM

We modelled expressive language function as predicted by language group, age at stroke group and its interaction, with timepoint variable as additional fixed effect as it did not increase fit as random effect. As shown in Table 3, language group showed significantly better outcomes for bilinguals over monolinguals (p=.02), while age at stroke group also has significant differences in a U-shape pattern. Once again, the first-year group showed a significant difference between language groups such that monolinguals had worse language expression outcomes than bilinguals at age 28 days to 1 year at stroke onset (Figure 1). Age at stroke group and its interaction with language group was a significant predictor of language expression PSOM (p=.01; Table 3).

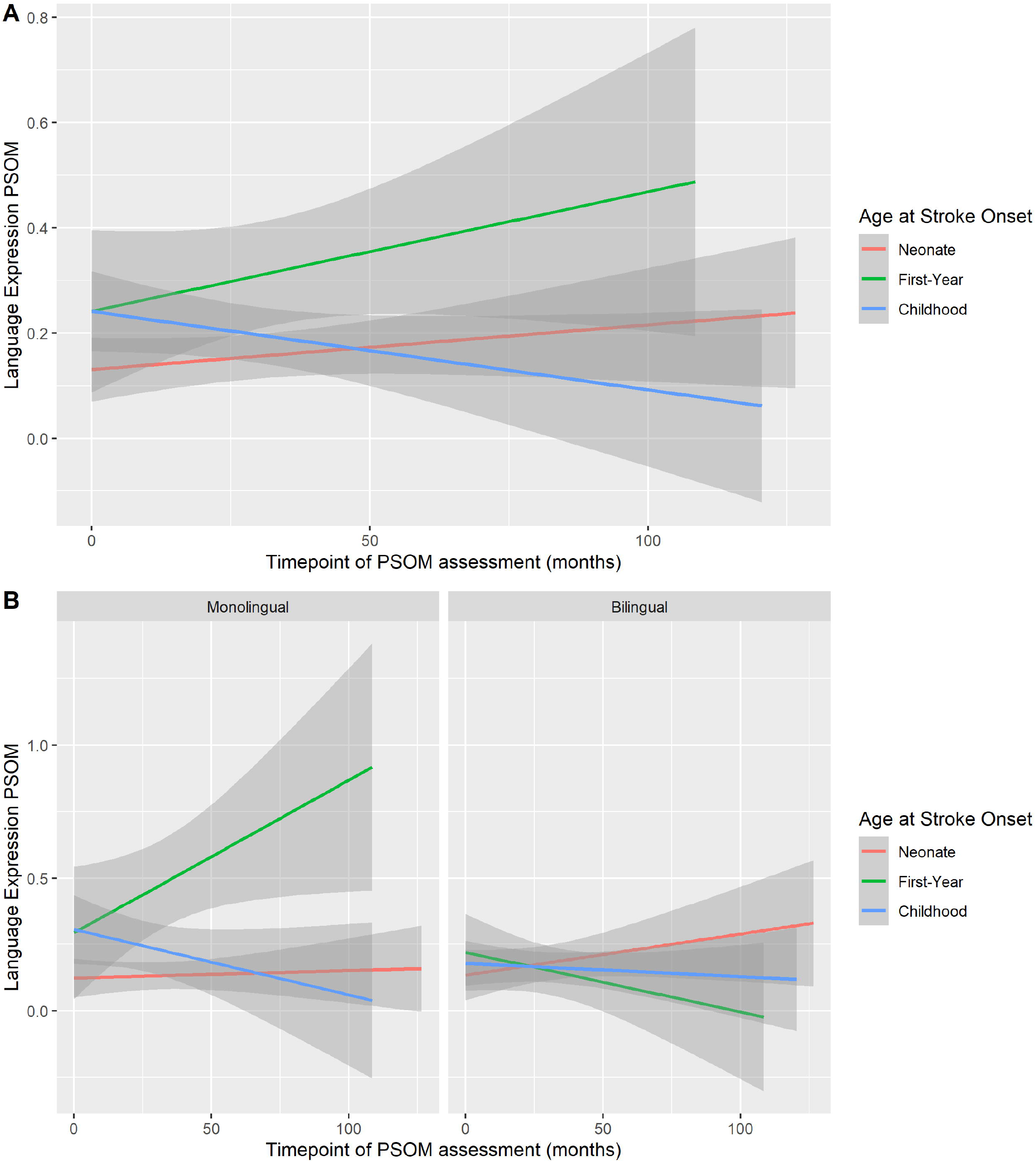

Using growth curve modelling (Figure 2A), we visualized trajectory differences for language expression PSOM scores over time by age at stroke onset groups. In Figure 2B, marked differences between the monolingual and bilingual trajectories particularly for the first-year group were evident. The monolingual group shows increases overtime if they had a stroke in their first-year, while the scores for bilingual group remained fairly steady over time.

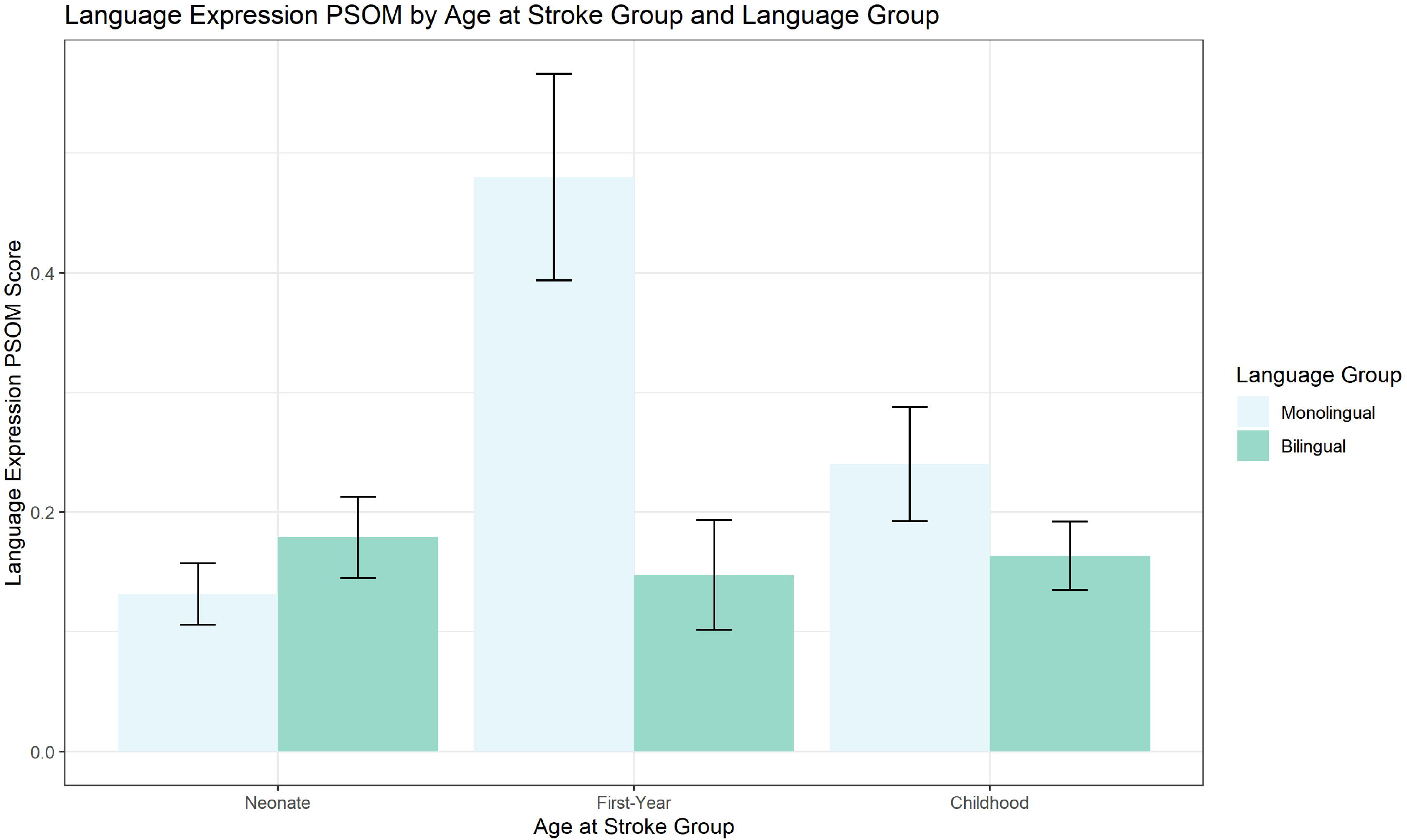

## Discussion

We evaluated the effects of bilingual exposure on the cognitive and linguistic development of children post-stroke, using the Pediatric Stroke Outcome Measure (PSOM) administered at several timepoints and up to 10.5 years post-stroke. First, we found no language group effect or interactions in the cognitive/behavioural PSOM model, suggesting that monolingual or bilingual status did not affect the cognitive/behavioural scores. Second, language group differences showed significantly better outcomes in the bilinguals in the case of language expression PSOM (p=.02) and combined language PSOM (p=.04). Third, better outcomes for bilinguals in the group that suffered stroke between 1 and 12 months of age was found in the language expression PSOM model (p=.01) and the combined (expression and comprehension) language PSOM (p=.04).

Age at stroke onset was a predictor of stroke outcome, such that the first-year group had the highest scores in all language PSOM subscales, consistent with previous findings^13^. Similarly, Trauner and colleagues found the trajectory of language development in monolingual children after perinatal stroke is significantly altered between the 1^st^ and 2^nd^ year of life compared to controls, such that by 2 years of age, their stroke group experienced delays in language production^33^. While this pertains to childhood stroke, it is possible that monolinguals of the first-year group showed similar outcomes for language expression, while the different outcome for bilinguals was reflective of the aforementioned linguistic advantage for bilinguals. We might also expect differences to be apparent if the stroke occurred outside of the neonatal period (28 days of birth), as this time frame would allow for sufficient bilingual exposure pre- and post**-**stroke. Outperforming monolinguals, bilingual children at this age make strides in language learning and metalinguistic skills^20^. Alternatively, at follow-up PSOMs, the first-year group could also be showing evidence of their language development, as they transition from a primarily home environment to more societal exposure to the language of assessment (English). Therefore, the difference in the language PSOM scores could equally be evidence of growth in English-language skills at this time in bilinguals (as the PSOM is administered in English), or evidence for a general advantage in language learning and metalinguistic skills in bilinguals in general which functions as a protective factor for pediatric patients.

The lack of an effect in the cognitive/behavioural model may suggest no difference between the two groups; alternatively, it may stem from the sensitivity of the PSOM in detecting changes in cognition. First, at this age, it is problematic to sharply distinguish cognition and language in these stroke-onset groups. Second, the agreement between normal/abnormal PSOM subscale scores was the weakest for the cognitive/behavioural subscale, compared to other PSOM subscales^25^. Finally, previous studies using the cognitive/behavioural PSOM have typically recruited older children aged 6 and onwards at study^10^. In other atypically developing populations, evidence for a cognitive advantage in bilinguals has been mixed. There has been a documented bilingual cognitive advantage in inhibition and switching in pediatric traumatic brain injury^34^ and in enhancing executive functioning on working memory tasks, but not mental flexibility and verbal fluency in children with epilepsy^35^. Given this, we cannot definitively conclude lack of the effect of bilingual exposure cognitive abilities. Future studies using finer-grain or more detailed cognitive measures with these populations to address this question further.

The timing of pediatric stroke during development entails complex cognitive and linguistic challenges in recovery^9^. Vulnerability during “sensitive periods” of development leave children open to the risk of further disruption in development, building on previous milestones^5^. Recovery has been conceptualized as categorical change on the PSOM severity classification system over time^13,26^; as assessed in the Recurrence and Recovery Questionnaire (based on the PSOM)^36^; and more generally, through a restoration/restitution of function or an adaptative compensation through substitution^37^. *Recovery* as it relates to post-stroke outcomes is predominantly used in the literature, though *development* is an intrinsically related concept. As such, conflation of these terms further complicate the situation of atypically-developing populations^37^.

## Conclusion

This is the first study evaluating the effect of bilingualism on language and cognitive development post-stroke. Our overall results indicate that patients from monolingual and bilingual environments follow similar development post-stroke when their cognitive abilities are considered, however, bilinguals had better language outcomes compared to monolinguals. Considering the limitations of the current study, more data is needed to make a more definitive conclusion about the cognitive development of monolingual and bilingual patients. Importantly, the current study did not find any negative effects of bilingual exposure on development in children post-stroke.

## Data Availability

All data produced in the present study are available upon reasonable request to the authors

https://dataverse.scholarsportal.info/dataverse/bilingualrecovery

## Notes

### Competing Interest Statement

The authors have declared no competing interest.

### Funding Statement

This study was funded by the Natural Sciences and Engineering Grant (RGPIN-2019-06523) to M.M and the Natural Sciences and Engineering Research Council - Alexander Graham Bell Canada Graduate Scholarship-Master's to K.I.L.

### Author Declarations

Ethics committee/IRB of The Hospital for Sick Children gave ethical approval for this work

### Summary of Updates

Analyses have been changed based on the grouping of bilinguals. More detail has been provided regarding the outcome measure. Tables and Figures have been revised.

