## Supplementary Materials 1 for "The Effect of Bilingual Exposure on Language and Cognitive Development in Children Following Ischemic Stroke" (from another version of this preprint: version 1, 2021-10-28)

**SUPPLEMENTARY MATERIALS: A**

Stratification of stroke-onset group variable was motivated by previous literature using the PSOM that indicated worse outcomes for those aged 1-12mos at stroke (Felling et al., 2020). Childhood stroke was subdivided into two groups: First-Year (1-12mos at stroke) and Childhood (13mos+). The following table provides the distribution for age of stroke onset:

|  | Perinate | First-Year | Childhood | Total |
| --- | --- | --- | --- | --- |
| Patients, n | 105 | 33 | 99 | 237 |

A test of normality showed Total PSOM, our dependent variable, was not normal. Hence, we chose the non-parametric Kruskal-Wallis test to compare the three groups. Results are seen in the table below. No other differences were found for the other dependent variables of interest (combined language PSOM, *p*=0.140; language expression PSOM, *p*=0.142; language reception PSOM, p=0.145; cognitive/behavioural PSOM, *p*=0.164).

|  | *Statistic* | *df* | *p* |
| --- | --- | --- | --- |
| *Total PSOM* | 12.156 | 2 | 0.002 |
| *Simple Effects* |  |  |  |
| Perinate – First-Year |  |  | 0.115 |
| Perinate – Childhood |  |  | 0.004 |
| First-Year – Childhood |  |  | 0.001 |
