## Supplementary Materials 2 for "The Effect of Bilingual Exposure on Language and Cognitive Development in Children Following Ischemic Stroke" (from another version of this preprint: version 1, 2021-10-28)

**B.** Summary of the linear mixed effects model with Cognitive/Behavioural PSOM as the dependent variable. Timepoint was retained as a random slope. Variables listed in the first column are independent variables. R^2^ marginal = 0.167, R^2^ conditional = 0.502.

Abbreviations: **^t^**, trend

|  | | *F* | | *Num df* | | *Den df* | | *P* | |
| --- | --- | --- | --- | --- | --- | --- | --- | --- | --- |
| Timepoint |  | 14.882 |  | 1 |  | 110 |  | <.001 | *** |
| Age at stroke group |  | 16.690 |  | 2 |  | 177 |  | <.001 | *** |
| Language Group |  | 0.2425 |  | 1 |  | 178 |  | 0.623 |  |
| Laterality |  | 2.7432 |  | 2 |  | 176 |  | 0.067 | ^t^ |
| Sex |  | 0.0272 |  | 1 |  | 180 |  | 0.869 |  |
| Age at stroke group ✻ Language Group |  | 0.4035 |  | 2 |  | 177 |  | 0.669 |  |
| Age at stroke group ✻ Laterality |  | 0.5271 |  | 4 |  | 175 |  | 0.716 |  |
| Language Group ✻ Laterality |  | 0.0888 |  | 2 |  | 185 |  | 0.915 |  |
| Age at stroke group ✻ Sex |  | 0.1140 |  | 2 |  | 176 |  | 0.892 |  |
| Language Group ✻ Sex |  | 0.4688 |  | 1 |  | 181 |  | 0.494 |  |
| Laterality ✻ Sex |  | 0.0528 |  | 2 |  | 183 |  | 0.949 |  |
| Timepoint ✻ Age at stroke group |  | 2.4719 |  | 2 |  | 104 |  | 0.089 | ^t^ |
| Timepoint ✻ Language Group |  | 0.2743 |  | 1 |  | 107 |  | 0.602 |  |
| Timepoint ✻ Laterality |  | 1.4965 |  | 2 |  | 112 |  | 0.228 |  |
| Timepoint ✻ Sex |  | 0.9272 |  | 1 |  | 106 |  | 0.338 |  |

**C.** Summary of the linear mixed effects model with Combined Language Subscales PSOM as the dependent variable. Variables listed in the first column are independent variables. R^2^ marginal = 0.088, R^2^ conditional = 0.345.

|  | | *F* | | *Num df* | | *Den df* | | *P* | |
| --- | --- | --- | --- | --- | --- | --- | --- | --- | --- |
| Timepoint |  | 0.1743 |  | 1 |  | 665 |  | 0.676 |  |
| Age at stroke group |  | 2.2922 |  | 2 |  | 151 |  | 0.105 |  |
| Language Group |  | 0.0116 |  | 1 |  | 151 |  | 0.914 |  |
| Laterality |  | 6.0632 |  | 2 |  | 150 |  | 0.003 | * |
| Sex |  | 0.4127 |  | 1 |  | 154 |  | 0.522 |  |
| Age at stroke group ✻ Language Group |  | 1.4739 |  | 2 |  | 152 |  | 0.232 |  |
| Age at stroke group ✻ Laterality |  | 1.0980 |  | 4 |  | 152 |  | 0.360 |  |
| Language Group ✻ Laterality |  | 0.1264 |  | 2 |  | 162 |  | 0.881 |  |
| Age at stroke group ✻ Sex |  | 0.1722 |  | 2 |  | 152 |  | 0.842 |  |
| Language Group ✻ Sex |  | 1.3390 |  | 1 |  | 156 |  | 0.249 |  |
| Laterality ✻ Sex |  | 0.3514 |  | 2 |  | 158 |  | 0.704 |  |
| Timepoint ✻ Age at stroke group |  | 1.9042 |  | 2 |  | 655 |  | 0.150 |  |
| Timepoint ✻ Language Group |  | 1.0654 |  | 1 |  | 670 |  | 0.302 |  |
| Timepoint ✻ Laterality |  | 0.5725 |  | 2 |  | 678 |  | 0.564 |  |
| Timepoint ✻ Sex |  | 0.1586 |  | 1 |  | 672 |  | 0.691 |  |

**D.** Summary of the linear mixed effects model with Language Expression PSOM as the dependent variable. Variables listed in the first column are independent variables. R^2^ marginal = 0.085, R^2^ conditional = 0.347.

Abbreviations: **^t^**, trend

|  | | *F* | | *Num df* | | *Den df* | | *P* | |
| --- | --- | --- | --- | --- | --- | --- | --- | --- | --- |
| Timepoint |  | 0.0250 |  | 1 |  | 663 |  | 0.874 |  |
| Age at stroke group |  | 1.5229 |  | 2 |  | 159 |  | 0.221 |  |
| Language Group |  | 0.0676 |  | 1 |  | 160 |  | 0.795 |  |
| Laterality |  | 4.9908 |  | 2 |  | 158 |  | 0.008 | * |
| Sex |  | 0.1415 |  | 1 |  | 163 |  | 0.707 |  |
| Age at stroke group ✻ Language Group |  | 2.5488 |  | 2 |  | 159 |  | 0.081 | ^t^ |
| Age at stroke group ✻ Laterality |  | 0.6539 |  | 4 |  | 159 |  | 0.625 |  |
| Language Group ✻ Laterality |  | 0.1971 |  | 2 |  | 168 |  | 0.821 |  |
| Age at stroke group ✻ Sex |  | 0.2068 |  | 2 |  | 159 |  | 0.813 |  |
| Language Group ✻ Sex |  | 1.6500 |  | 1 |  | 162 |  | 0.201 |  |
| Laterality ✻ Sex |  | 0.4755 |  | 2 |  | 164 |  | 0.622 |  |
| Timepoint ✻ Age at stroke group |  | 1.9998 |  | 2 |  | 654 |  | 0.136 |  |
| Timepoint ✻ Language Group |  | 1.5138 |  | 1 |  | 668 |  | 0.219 |  |
| Timepoint ✻ Laterality |  | 0.8339 |  | 2 |  | 676 |  | 0.435 |  |
| Timepoint ✻ Sex |  | 0.5164 |  | 1 |  | 670 |  | 0.473 |  |

**E.** Summary of the linear mixed effects model with Language Reception PSOM as the dependent variable. Variables listed in the first column are independent variables. R^2^ marginal = 0.081, R^2^ conditional = 0.265

|  | | *F* | | *Num df* | | *Den df* | | *P* | |
| --- | --- | --- | --- | --- | --- | --- | --- | --- | --- |
| Timepoint |  | 0.48775 |  | 1 |  | 665 |  | 0.485 |  |
| Age at stroke group |  | 3.48946 |  | 2 |  | 141 |  | 0.033 | * |
| Language Group |  | 0.24015 |  | 1 |  | 139 |  | 0.625 |  |
| Laterality |  | 7.31494 |  | 2 |  | 138 |  | <.001 | *** |
| Sex |  | 0.39426 |  | 1 |  | 146 |  | 0.531 |  |
| Age at stroke group ✻ Language Group |  | 0.40322 |  | 2 |  | 139 |  | 0.669 |  |
| Age at stroke group ✻ Laterality |  | 1.71086 |  | 4 |  | 140 |  | 0.151 |  |
| Language Group ✻ Laterality |  | 0.15806 |  | 2 |  | 151 |  | 0.854 |  |
| Age at stroke group ✻ Sex |  | 0.25585 |  | 2 |  | 142 |  | 0.775 |  |
| Language Group ✻ Sex |  | 0.67113 |  | 1 |  | 146 |  | 0.414 |  |
| Laterality ✻ Sex |  | 0.27795 |  | 2 |  | 148 |  | 0.758 |  |
| Timepoint ✻ Age at stroke group |  | 1.84141 |  | 2 |  | 660 |  | 0.159 |  |
| Timepoint ✻ Language Group |  | 0.38061 |  | 1 |  | 664 |  | 0.537 |  |
| Timepoint ✻ Laterality |  | 0.25050 |  | 2 |  | 673 |  | 0.778 |  |
| Timepoint ✻ Sex |  | 0.00283 |  | 1 |  | 674 |  | 0.958 |  |
